# Data-Driven Inference of COVID-19 Clinical Prognosis

**DOI:** 10.1101/2020.08.27.20183202

**Authors:** Joaquín Salas, Dagoberto Pulido, Omar Montoya, Isaac Ruiz

## Abstract

Knowing the most likely clinical prognosis for a patient infected with SARS-Cov-2 could offer guidelines for tracking their medical evolution, improving attention, and assigning resources. Aiming to assess a patient’s status quantitatively, we explore the analysis of existing clinical information using data-driven methods. Our goal is to extract the characteristics distinguishing between those COVID-19 patients that improve and those who die. In our approach, we select the relevant features using the algorithm of Boruta, a wrapper framework that takes input from classifiers generating relevance assessment of the predictors. Using the extracted features, we train machine learning classifiers, including Random Forests, Support Vector Machine, Extreme Gradient Boosting, and Neural Networks. We assess the performance of the classifiers using Precision-Recall and ROC analysis, establishing the ranges at which risk assessment permits effective decision-making. Our research highlights that local regions present unique sets of essential features, that it is possible to construct effective classifiers based on clinical data, and that an ensemble of classifiers results in the best performing discriminant.

## Introduction

COVID-19 is an infectious disease caused by the SARS-CoV-2 virus. Some common symptoms at the beginning of a COVID-19 infection are fever, cough, and fatigue. Other signs that may appear afterward include sputum production, myalgia, headache, hemoptysis, diarrhea, dyspnea, and lymphopenia^1^. However, currently, the definitive diagnosis of COVID-19 is made by quantitative reverse-transcription polymerase chain reaction (qRT-PCR) assays, which are molecular techniques that consist of obtaining a large number of copies of a nucleic acid fragment. Even though clinical diagnosis^1^ and molecular techniques^2^ allow advancing in the detection of infected patients, once a physician declares a case confirmed positive, the patient evolution will determine whether the patient improves or dies. The quantitative inference of the likely outcome permits to modify medical treatment, administer resources, and proactively define courses of action. In this paper, we propose a data-driven approach to track symptomatology and assess a confirmed positive COVID-19 patient’s clinical prognosis.

As the amount of information available increases, machine learning emerges as an alternative that provides supplemental information to infer outcomes based on predictors. Its response speed is its strength in the presence of exponential growth in the number of infections. In this framework, the machine learning community has been developing methodologies allowing to predict the change in the rate of infection over time^3^, search for substances suitable for treatment^4^, contact tracing to track the spread of the infectious^5^, and classify genome relevant for COVID-19^6^. Recent results highlight the importance of large datasets in extracting granular risk factors associated with COVID-19^7^. In this regard, the use of non-linear classifiers permits to capture complex relations not evident on a predictor by predictor basis. In this document, we explore the relationship among factors to predict the outcome of COVID-19 positive cases. Besides confirming previous research^8,9^ claiming that it is possible to solve such a task with a high level of confidence, our contributions include the following:

- We show that the characteristics predicting COVID-19 clinical prognosis have a substantial local component, highlighting the need to treat regions differently.
- We show that an ensemble of classifiers outperforms individual classifiers for the COVID-19 clinical prognosis task.
- We make publicly available our code for other research to corroborate our claims and as a stepping stone for further inquiries.

We structure the rest of the paper as follows. In the next section, we contextualize our research based on the current literature related to COVID-19. Then, we detail the strategy we have developed to extract the essential features of our problem. Afterward, we proceed to describe the classifiers employed in our analysis to predict patients’ clinical prognosis based on their clinical records. After presenting and discussing our results, we conclude summarizing and delineating future lines of enquiring.

## Related Literature

In our approach, we build an ensemble of machine learning classifiers to identify the risk of decease in COVID-19 patients. To achieve this, we first select what features are more relevant to distinguish patients between surviving and dying. In this section, we describe the literature related to these topics.

Lalmuanawna *et al*.^10^ present a review of applications of Machine Learning (ML) and Artificial Intelligence (AI) for COVID-19. They perceive that most models are not deployed enough to show their real-world operation, although there is ample space for them to impact the pandemic. For instance, Sharma^11^ employ Convolutional Neural Networks (CNN) to infer the results of quantitative reverse-transcription polymerase chain reaction (qRT-PCR) molecular test out of lung Computer Tomographies (CT) scans, employing 618 images of healthy people (175), patients with influenza-A pneumonia (224) and patients infected with COVID-19 (219 images). Similarly, Mei *et al*.^12^ explore the use of AI techniques for rapid diagnosis for COVID-19 as an alternative to PRC testing. They employ CT images, clinical symptoms, exposure history, and laboratory testing to diagnose patients who are positive for COVID-19. To analyze the images, they use CNN while analyzing the clinical data; they employ a mixture of Support Vector Machine (SVM), Random Forest (RF), and Multi-Layer Perceptrons (MLP). Although their research focuses on diagnosis while we are interested in prognosis, we coincide in some of the ML techniques to construct the classifiers and voting schemes.

Closer to our problem, researchers have employed survival analysis to distinguish patients between general, severely ill, and deceased. That is the case of Yan *et al*.^13^, who present an ML method to infer clinical outcome based on the use of three blood biomarkers: Lactic dehydrogenase (LDH, associated with tissue breakdown in pulmonary disorders), lymphocyte (a subtype of white blood cell essential part of the innate immune system), and high-sensitivity C-reactive protein (hs-CRP, which increases in the blood during inflammation and infection processes). By collecting samples from 485 patients, they can issue a ten-day prognosis forecast with 90% accuracy employing an Extreme Gradient Boosting (XGB) classifier. In their approach, they train with 262 samples, validated with 113 cases, and tested on 29 achieving near-perfect performance. In our research, we show that it is possible to achieve a similar performance level using other clinical symptoms (see Table 2), perhaps at the expense of requiring more data samples. Also, practitioners have developed techniques for risk factor selection and outcome prediction. For instance, Nemati *et al*.^14^ implement survival analysis methods on 1,182 COVID-19 patients to predict recovery and discharge time of hospitalized patients employing age and sex features. They obtain the best results with XGB. Schwab *et al*.^15^ implement ML algorithms to predict the outcome of a SAR-CoV-2 test and whether a positive patient will require hospitalization or intensive care. They use clinical and blood analysis data from 5,644 patients. Similarly, we perform an analysis to determine which clinical features are predictive, and to what extent, for each of the aforementioned clinical tasks.

In a large scale experiment, Pourhomayoun and Shakibi^9^ assess mortality risk in patients with COVID-19 using a dataset of 117,000 patients from 76 countries. They train several ML algorithms, including SVM, Artificial Neural Networks (ANN), RF, Logistic Regression (LR), and k-Nearest Neighbors (kNN), being ANN the best performer. Starting with 117 features, from a public domain dataset, they end up with 42. Differently, we employ a wrapper feature selection method, but more importantly, we emphasize the role of regional data in the extraction of essential features. Hojo *et al*.^16^ also propose to predict the COVID-19 outcome using ML techniques. They apply an array of ML methods, including LR, Linear Discriminator Analysis (LDA), Naive Bayes (NB), kNN, Decision Trees (DT), XGB, and SVM to 13,690 closed cases patients, where the number of deaths corresponds to 8.21% and 6.11% in the training and validation sets. They solve this unbalanced dataset employing oversampling and achieve Receiving Operating Characteristics (ROC) Area Under the Curve (AUC) of 0.92 with Precision-Recall (PR) AUC of 0.5 in their best model. Although our dataset has not the same level of unbalancing, we instead employ undersampling combined with cross-validation to avoid dropping information. Bertsimas *et al*.^17^ develop an assessment of mortality risk using a data set obtained in an international multi-center effort focused on Europe and the US. Their classifier, based on XGB, employs clinical and laboratory predictors. Among the most critical factors for mortality risk, they obtain age, decrease oxygen saturation, elevated levels of hsCRP, blood urea nitrogen, blood creatinine, and blood glucose.

Our review of the literature shows that there is an opportunity to apply wrapper based techniques for feature selection and ensemble of classifiers to predict clinical prognosis in COVID-19 patients. Furthermore, an exciting aspect of our research is that one may use the data resulting from medical interviews to achieve state-of-the-art performance, emphasizing the analysis of regional conditions to improve tracking of the important features.

## Important Features Extraction

Given the database, in our study, we carry out an analysis to select the variables that best distinguish between improved and deceased COVID-19 patients. With this aim, we use the implementation of the Boruta algorithm^18^, a wrapper-type method that accepts variable importance measure methods as underlying classifiers. Many other selection methods consider all the characteristics simultaneously, such as Mutual Information and Super Casual Correlation^19^. However, we selected Boruta’s algorithm because it considers multiple associations simultaneously, without an exhaustive search. This strategy is a departure from similar approaches which are based on data correlation between predictors^9^.

Boruta’s algorithm begins by defining shadow variables for each predictor. Then, the method shuffles the shadow variable’s values over the observations of the same predictor. Boruta’s algorithm compares the relative importance of the original variables and the shadow variables. If the importance value of an original variable is statistically higher or lower than the maximum importance of the best-valued shadow variable, the method labels the predictor as important or not important. Then, Boruta discards the tagged variables and repeats the procedure until it tags all the variables.

One could employ the Boruta framework with classifiers delivering importance measures, such as RF, XGB, or Random Ferns^20^. The RF^21^ algorithm naturally handles a measure of importance. For a tree, a variable’s importance is related to the increase in precision achieved with the partitions made in the nodes, typically measured in terms of the Gini index^22^ or the entropy/information gain^23^. For a forest, the method averages the importance of a predictor over all trees. However, due to the RF algorithm’s haphazard nature, each run may result in a different selection. To attack this problem, we run the Boruta algorithm k times, recording the characteristics and relative importance assigned to them. With each of the resulting subsets of attributes, one could construct a classifier, remaining to identify which offers the best generalization capacity.

## Classifying Clinical Prognosis

Using the characteristics considered important, we built a set of classifiers using Neural Networks (NN), Random Forests (RF), Support Vector Machine (SVM), and Extreme Gradient Boosting (XGB).

*Neural Networks* are universal function approximators^24^ represented by layers of neurons, each having inputs and outputs. Each neuron receives a weighted sum of the inputs plus a bias. If we normalize the weights to unity, one may imagine the result being the distance from the inputs to the hyperplane defined by the weights, when one adds the bias. The result passes through a non-linear activation function. If we assume a sigmoid activation function, we may resemble the computation of a logistic regression classifier at each neuron and a function approximation to a possible non-linear boundary with the neurons of each layer^25^. The output of each layer will form the input to the next layer. In the last layer, we may have a single neuron with a sigmoid activation function, which output and complement will represent the probability of membership for the classes in our problem.

*Random Forests* ^21^ is an extension of Decision Trees where one trains a particular tree with a subset of the dataset available. The algorithm selects a random subset of the predictors at each split, and one creates a large number of trees, naturally constructing an ensemble of classifiers. Like other tree-based methods, Random Forests handle equally well regression and classification problems and are similarly well adapted to deal with numerical and categorical variables. Furthermore, the independence of the ensemble elements reduces the variance during voting for classification or averaging for regression.

*Support Vector Machines* ^26^ are constructed on the criterion of maximizing the distance between discriminatory hyperplanes and the classes. Once maximized the margin, the closer points at either side of the hyperplane are called support vectors. To allow misclassification, one introduces a cost budget C. In the general case, where there is no hyperplane dividing the classes, one projects the data over non-linear, possibly higher-dimensional spaces, where the expectation is that the data will be linearly separable.

*Extreme Gradient Boosting* ^27^ combines the approach of creating a large number of decision trees in Random Forest with the construction of a cascade of weak classifiers typical of boosting strategies ^28^. The approach incorporates sequential tree growing where, like in boosting, the weight is adjusted for the predictors wrongly classified by the previous tree. In turn, just like in Random Forest, each sequential tree is constructed with a random subsample of the data (in a process known as bagging) and a random subset of predictors during splitting.

**Table 1.** Datasets employed in our experiments. Dataset corresponding to the Mexican stated of Mexico City (DF), Sinaloa (SL) and Queretaro (QT).

|  | DF | SL | QT |
| --- | --- | --- | --- |
| Date | 2020.06.26 | 2020.07.30 | 2020.08.02 |
| Initial records | 46,834 | 21,761 | 5,812 |
| COVID-19 confirmed positive | 12,358 | 5,947 | 1,663 |
| Initial features | 61 | 61 | 61 |
| Important features | 21 | 25 | 13 |
| Records with clinical prognosis | 8,150 | 5,442 | 1,103 |
| Recovery | 5,028 (61.7%) | 4,187 (76.9%) | 841 (76.2%) |
| Death | 3,122 (38.3%) | 1,255 (23.1%) | 262 (23.8%) |

*Ensembles* are predictive models interpreting multiple classifiers^29^. Boosting and bagging strategies could also construct classifiers’ ensembles, but in our case, we aim to interpret it as a voting mechanism. We hypothesize that given that we construct the classifiers on a somewhat different criterion and that the feature space is large, each classifier will likely describe a somewhat different discrimination surface. Although one could pursue multiple strategies, we implement the voting mechanism as a supervised ensemble regression via a neural network. Our objective is to use the neural network as a general regression function *f* to map the output *p* = (*p*_xgb_, *p*_svm_, *p*_rf_, *p*_nn_) of the previous classifiers into a final prediction q. We fine-tuned the neural network employed for voting using a grid search on the number of layers and the number of neurons per layer settled with an architecture of one hidden layer with one unit. This procedure defined a hyper-plane

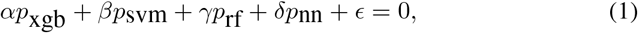

with each of the states defining a different set of parameters and where *p*Xgb, *p*_svm_, *p*_rf_ and *p*_nn_ correspond to the XGB, SVM, RF and NN classifiers output.

During the learning process, we split by half the data into training and validation sets. Furthermore, in each partition, we undersample the most abundant class to form the input dataset. We repeat this operation *l* times for each classifier, in a process known as cross-validation, each time performing partitions with the same policy, but on a random subset of the data. For each classifier, we evaluated performance using ROC (Receiving Operating Characteristics) curves and Precision-Recall. As a result of repeated performance evaluation, we have variations that we can summarize statistically using the mean and standard deviation to estimate its most likely value and corresponding uncertainty.

## Experimental Results

In our experiments, we aim to determine the ability of the feature selector to produce relevant descriptors and the usefulness of the data to permit reliable classifiers. To that end, we take a dataset of confirmed positive COVID-19 patients and process it to extract essential features and construct classifiers to evaluate ROC and Precision-Recall analysis.

**Figure 1.**
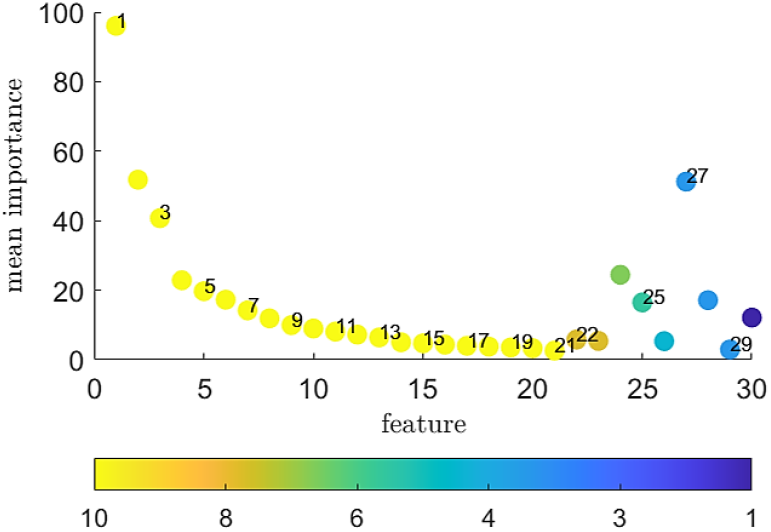
Feature Selection. Important characteristics in the process of discriminating between confirmed and discarded cases for COVID-19 using the Boruta algorithm. Here, we illustrate the Mexico City (DF) case. The horizontal and vertical axis represents the feature number and estimated importance. The color represents how many times the algorithm chooses a feature: In this example, yellow represents ten while deep blue represents one (best seen in color, see Table 2 for further details).

### Experimental Setup

For our study, we have three datasets containing records from anonymized records for the Mexican states of Mexico City (DF), Sinaloa (SL) and Queretaro (QT) provided by the Mexican Secretariat of Health, containing COVID-19 confirmed positive cases after a qRT-PCR test (see Table 1). Initially, a dataset record contains 192 variables. Some of these variables are associated with administrative information, including the registration number; post-diagnosis treatment of COVID-19, such as whether a hospital admitted the patient to the Intensive Care Unit (ICU); or personal information, such as name and address. As a pre-process, we remove these features to leave only those that would make up a diagnostic clinical verification list. After removing the related administrative features, the number of predictors was 61. We further split the dataset, keeping only the records with prognosis registering a patient recovery or decease.

The computer we use to process the information runs on the Windows 8.1 operating system and consists of a 64-bit Intel i7-3770 processor at a clock frequency of 3.4Hz, and operating with 16 GB of RAM. We developed our computer programs in **R** version 3.6.3.

### Feature Selection

To extract the important features, we ran Boruta’s algorithm ten times for each of our datasets, leaving 21, 25, and 13 characteristics deemed necessary for DF, SL, and QT, respectively (see Figure 1 and Table 2). These features appeared in each iteration of Boruta. We removed those features that did not result in each iteration, and those deemed unimportant by Boruta. It is worth noting that by far, age is the most important feature. Also, please note that each state has a different number of features, highlighting the importance of performing regional analysis of the information. However, note that while the number of features changes, their relative order does not, *i.e*., the characteristics common to different states preserve their relative order.

**Table 2.**
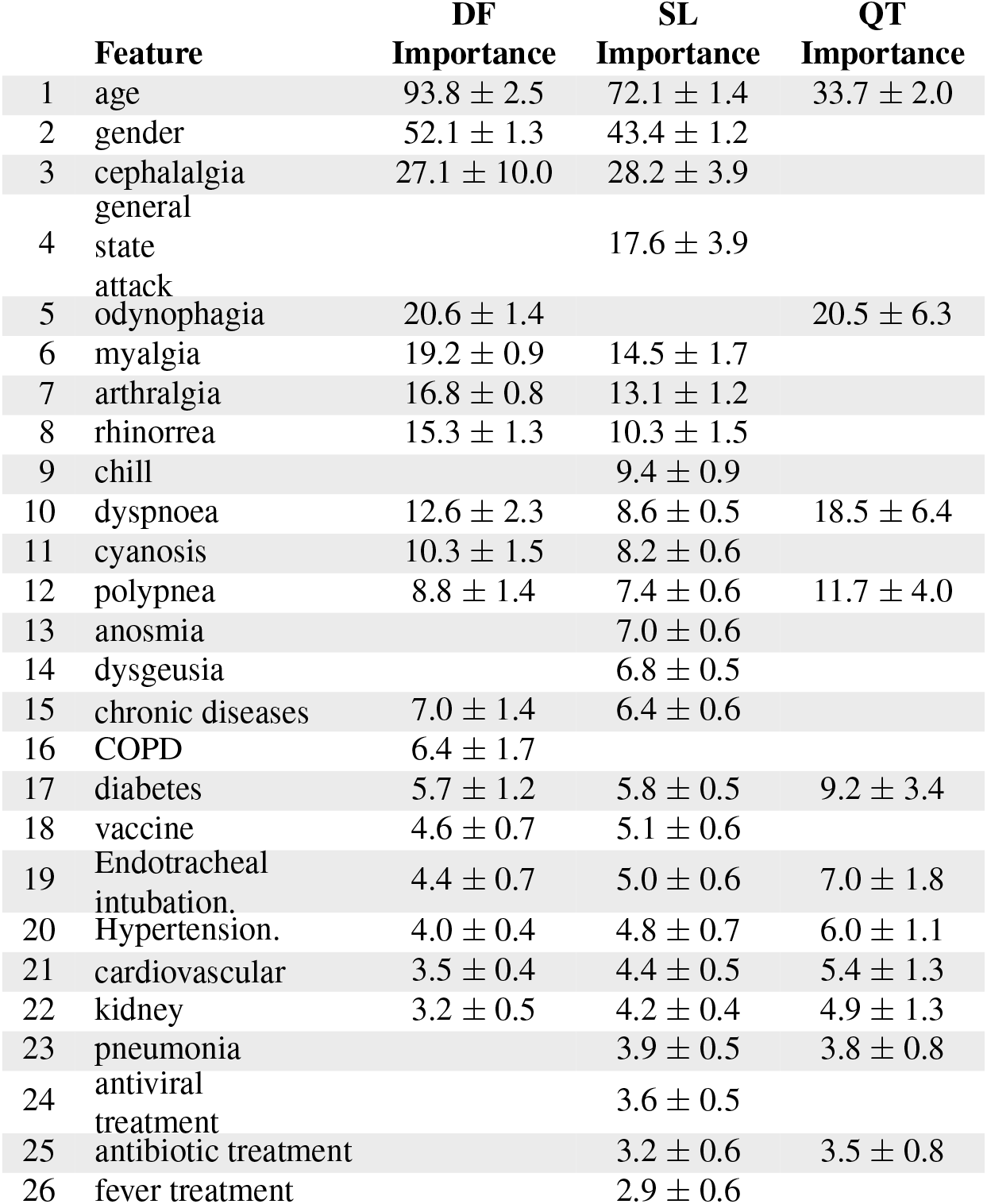
Feature importance. We show the variables deemed important for Mexico City (DF), Sinaloa (SL), and Queretaro (QT). The importance is the mean difference, across the trees in a forest, of votes cast for the correct class in the original predictor m and the same predictor with the values permuted.

**Figure 2.**
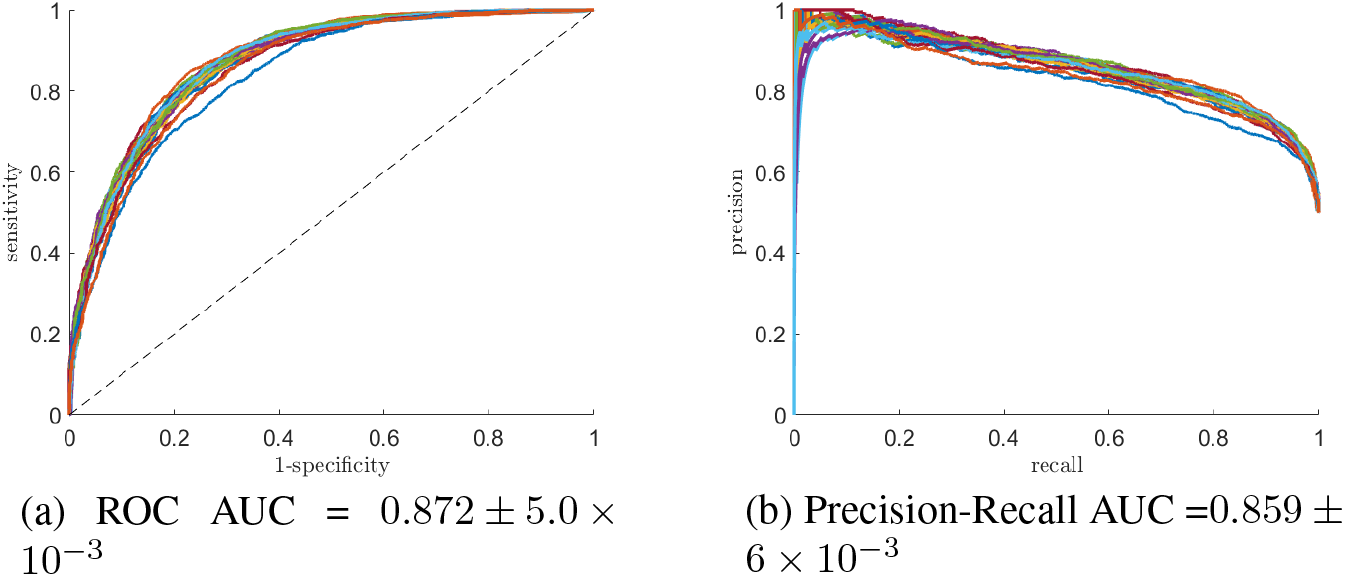
Evaluation of performance. The ROC and Precision-Recall curves summarize the resulting elements in the confusion table at different threshold values. Here, we illustrate the curves for the case of Mexico City (DF) for 30 random partitions for the dataset’s training and validation. We provide uncertainty at one standard deviation.

### Classifiers’ Fine-Tuning

For each set of features deemed important for a given state, one could construct classifiers, remaining to identify their performance measured as their generalization capacity. To do this, we first fine-tune the hyperparameters for each of the models. In the case of RF, we refined the hyperparameters corresponding to the number of variables randomly sampled at each split and the number of trees in the random forest. We use ten-fold cross-validation, for the former, and grid search, for the latter, resulting in 4 (four) for the number of variables and 1,500 for the number of trees. For the SVM, we fine-tuned the cost to 128 using 10-fold cross-validation and testing 10 (ten) different cost values. In the case of the XGB algorithm, it requires considerably more hyperparameters to tune. We fine-tuned using a random search on the learning parameter (0.148), the fraction of predictors used for splitting at each level (0.341), the maximum depth for the trees (10), the fraction of the data to sample (0.453), the minimum number of observations in a terminal node (1), the minimum reduction in the loss function to grow a new node in the tree (0.149) and the number of trees (696). In the case of the neural network-based classifier, we fine-tuned the number of hidden layers and the number of units within each layer. Interestingly, the best performer consisted of one layer and one neuron in that layer. One may interpret this result as a form of logistic regression where the inputs, weights, and bias define the hyperplane dividing classes. The sigmoid activation function assigns values between 0 (zero) and 1 (one) to the class membership. Finally, we also fine-tuned the neural network employed for voting using grid search and settling with an architecture of one hidden layer with one unit (see Figure 3).

### Classifiers’ Training and Performance Evaluation

Using these hyperparameters and the characteristics considered significant, we built classifiers based on RF, SVM, XGB, NN, and the final ensemble. For learning, we split the data into 50% for training and 50% for validation. Furthermore, before training or testing, we undersample the most abundant class to make it contain the same number of registers as the smaller one. We repeat this process 30 times, each performing a partition with the same percentage, but using uniform random sampling. Next, for the classifier, we evaluated its performance using ROC and Precision-Recall curves. Interestingly, taken one by one, the simple NN was the best performer, followed by the XGB, SVM, and RF, in that order of performance. Nonetheless, the overall best performer consisted of grouping all the classifiers and weighting their contribution. As a result of repeated performance evaluation, we have variations that we summarize in Table 3 using the mean and uncertainty at one standard deviation. The results for each state change, but are above 0.85 in the ROC and Precision-Recall AUC. In Figure 2, we illustrate the resulting performance curves for Mexico City.

**Figure 3.**
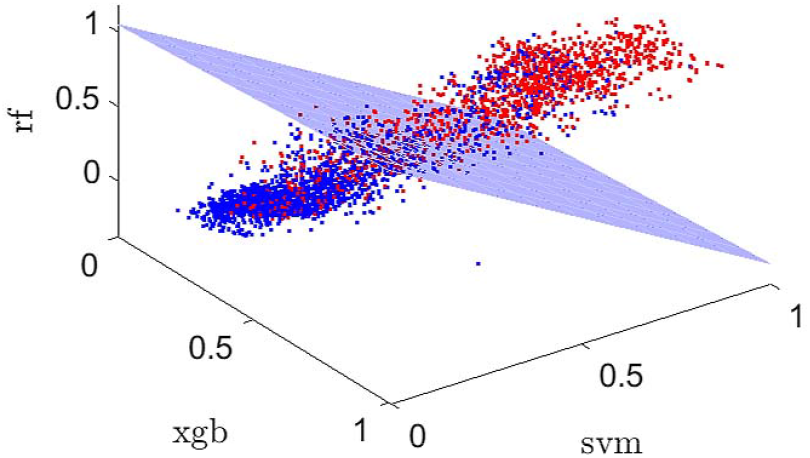
Classifiers Voting. Illustration of the hyperplane defined by the ensemble for three out of the four classifiers. Please note that the classifier we employed used four classifiers. The neural networks classifier is not illustrated in this figure.

**Table 3.** Performance result for the RF, SVM and the XGB classifiers. We represent the mean AUC value for the 30 iterations where the training and validation dataset splits have been chosen at random. We express uncertainty at one standard deviation

|  |  | XGB | SVM | RF | NN | ensemble |
| --- | --- | --- | --- | --- | --- | --- |
| DF | ROC | 0.84±0.01 | 0.83±0.01 | 0.82±0.01 | 0.84±0.01 | **0.87±0.01** |
|  | PR | 0.82±0.01 | 0.79±0.01 | 0.78±0.01 | 0.82±0.01 | **0.86±0.01** |
| SL | ROC | 0.89±0.01 | 0.89±0.01 | 0.89±0.01 | 0.90±0.01 | **0.93±0.08** |
|  | PR | 0.87±0.01 | 0.86±0.01 | 0.85±0.01 | 0.88±0.01 | **0.91±0.01** |
| QT | ROC | 0.85±0.03 | 0.85±0.02 | 0.84±0.02 | 0.85 ± 0.04 | **0.89±0.02** |
|  | PR | 0.82±0.03 | 0.82±0.03 | 0.80±0.04 | 0.81 ± 0.05 | **0.85±0.04** |

An intriguing question is the interpretation of the results in Table 3. The ROC analysis defines class separability, whereas PR analysis aims to characterize the class of interest^30^. Whereas the former works best with balanced classes, the latter, by not considering the true negatives, is more consistent with unbalanced datasets^31^. In our case, we mind making the classes balanced by design and iterating using cross-validations. We believe that this experimental setup will offer confidence in the framework’s capability to generalize well for unseen data. Overall, the ensemble classifier has a positive effect in performance.

**Table 4.**
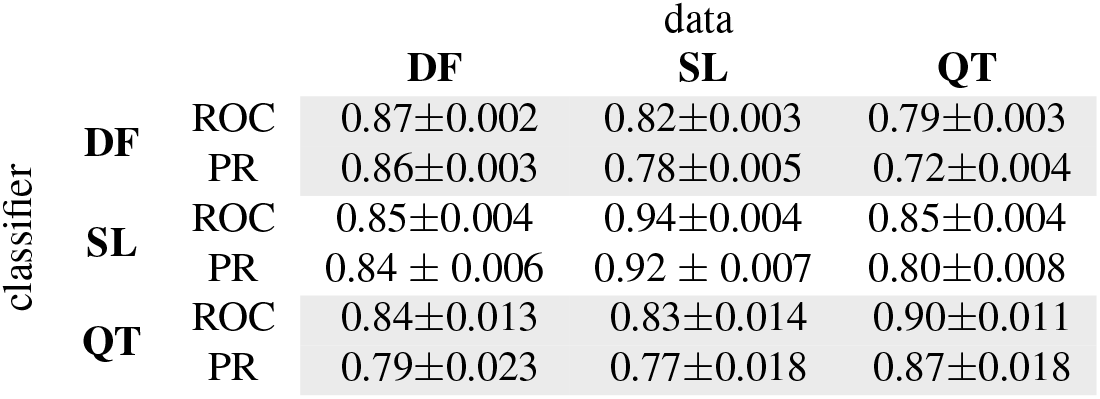
Performance with unseen data. The classifiers (rows), trained with data from one state, are tested with data from other states (columns). DF, SL, and QT stand for Mexico City, Sinaloa, and Queretaro, respectively. Rows correspond to ROC/Precision-Recall AUC mean value plus/minus one standard deviation.

Another interesting question to explore is the capability of the classifiers to discriminate unseen data. To that effect, we employed the classifier trained with the data from one state and utilized it with data from another state. Since the features that best discriminate clinical prognosis are different for each state, we have to utilize only the features required by the classifier. Table 4 shows the results. The rows represent the state employed to construct the classifier while the columns represent the state origin of the data. In this experiment, we used all the records in the dataset. As expected, we obtained the best performance when the classifier and the data corresponded to the same state. Interestingly, the classifiers keep a considerable inference capability.

## Conclusion

Through a classifier, one establishes the mapping between input characteristics and an output class. Given a confirmed positive case to COVID-19, this document shows that it is feasible to construct a high-performance classifier to predict a patient’s clinical prognosis. Although characteristics associated with COVID-19 are now widely known^8^, our research quantifies the importance of each of them related to the patient clinical prognosis. Likewise, the classifier presented here offers the ability to assign a measure of certainty regarding the prediction. A decision-maker may set a threshold for assignment to one or the other class balancing risks and costs. Decision-makers may play out these tradeoffs using the performance curves as they permit to know the decision thresholds that allow assigning an outcome to a patient with COVD-19. Our tool can thus have a critical complementary value for patients’ follow-up.

During a fast-paced epidemy, such as COVID-19, it is essential to have unbiased assessments for decision-making. This research shows that a data-driven approach, within the context of the dataset used for training, may offer an alternative that is fast, inexpensive, and error-bound.

## Data Availability

The code and data has been made public on github

https://github.com/joaquinsalas/COVID19-DataDriven-Classifier

## Data Dictionary

The features resulted after our analysis to identify essential features include:

1. `Age`. Age of the patient in years.
2. `Gender`. Self-declared patients’ gender.
3. `Cephalalgia`. Headache.
4. `General state attack`. Poor general condition.
5. `Odynophagia`. Pain while swallowing.
6. `Myalgia`. Muscle pain.
7. `Arthralgia`. Joint pain.
8. `Rhinorrea`. Free discharge of nasal mucus fluid.
9. `Chill`. A feeling of coldness during high fevers.
10. `Dyspnoea`. Shortness of breath.
11. `Cyanosis`. Bluish coloration due to low oxygen saturation in the skin.
12. `Polypnea`. Deep and rapid breathing.
13. `Anosmia`. Lack of ability to detect smells.
14. `Dysgeusia`. Distortion of the sense of taste.
15. `Chronic diseases`. A chronic condition in the patient’s health.
16. `COPD`. Chronic Obstructive Pulmonary Disease.
17. `Diabetes`. High level of blood sugar over a prolonged period.
18. `Vaccine`. Whether the patient received a vaccination.
19. `Endotracheal intubation`. Endotracheal intubation. The physicians place a tube into the trachea through the mouth or nose.
20. `Hypertension`. Long-term medical persistent condition in which the patient’s blood pressure in the arteries is elevated.
21. `Cardiovascular`. Heart condition manifested by diseased blood vessels, structural problems, and blood clots.
22. `Kidney`. The kidneys lose their ability to remove waste and balance fluids.
23. `Pneumonia`. The air sacs in one or both lungs are filled with fluid or pus, possibly causing respiratory difficulties, including cough with pus, fever, and chills.
24. `Antiviral treatment`. Whether a drug has been received for treating viral infections.
25. `Antibiotic treatment`. Whether a drug has been received for treating bacterial infections.
26. `Fever treatment`. Whether a plan has been followed to reduce fever.

## Acknowledgements

Thanks to the Mexican Health Ministry for the dataset on top of which we developed this research. Thanks to Alejandro Gomez for his support implementing the COVID-19 clinical prognosis calculator. Dagoberto Pulido and Omar Montoya received scholarships from CONACYT. This work was partially funded by SIP-IPN 20201357 for Joaquín Salas.

## Author biography

Joaquín Salas is a professor in the field of Computer Vision at Instituto Politécnico Nacional. Member of the Mexican National Research System, his research interests include the monitoring of natural systems using visual perception and aerial platforms. Salas received a Ph.D. in computer science from ITESM, México. He has been a visiting scholar at Stanford University, Duke University, Oregon State University, Xerox PARC, the Computer Vision Center, and the École Nationale Supérieure des Télécommunications de Bretagne. He has served as co-chairperson of the Mexican Conference for Patter Recognition three times. Salas was Fulbright scholar for the US State Department. He has been invited editor for Elsevier Pattern Recognition and Pattern Recognition Letters. For his services at the Instituto Politecnico Nacional, he received the *Lázaro Cárdenas* medal from the President of Mexico.

Omar Montoya is a PhD student at Instituto Politecnico Nacional (IPN) in the area of Image Analysis, his research interests include multi-modal perception systems for robotic applications. Montoya received a Master’s degree in Automation from the Instituto Tecnologico de Queretaro, Mexico Dagoberto Pulido is a Ph.D. student at Instituto Politécnico Nacional (IPN). His research interests include computer vision, remote sensing, and data analysis. Pulido received a Master’s in Advanced Technology degree from IPN, México.

José Angel Isaac Ruiz Mata. A surgeon from the Universidad Auténoma Metropolitana, Mexico, with a Master in Public Health with a Health Services Administration specialty area. He has served as President of the Board of Directors of the First Health Cooperative in Mexico, Coordination of Information, Planning, and Evaluation for the Comprehensive Nutrition Program with the care of more than 100 thousand children under five years of age on prevention and care of child malnutrition. Medical Coordinator in the General Directorate of Health Promotion participated in the design of indicators for evaluating the Vete Sano, Regresa Sano Program of attention to migrants in Mexico. Epidemiologist of a hospital and family medicine unit for the epidemiological surveillance of communicable and non-communicable diseases, as well as infections associated with health care, as an advisor to the General Directorate of Health Promotion, generated the National Strategy for the Prevention and Control of Diabetes, Overweight and Obesity for the 2013-2018 term. Currently as an Epidemiologist in a Regional General Hospital in IMSS Querétaro. Professor of the specialty in preventive medicine and public health at the National Institute of Public Health (INSP) and master’s degrees in Public Health of the INSP, health services management of the UVM, in migratory studies of the Ibero-American University and international cooperation of the Institute Mora of CONACYT. Currently a doctorate in the Senior Management in Health Establishments program.

## Declaration of conflicting interests

The authors declared no potential conflicts of interest with respect to the research, authorship, and/or publication of this article.

## Funding

This work was partially funded by SIP-IPN 20201357.

## Supplemental material

To foster further research, allowing other researchers to verify our results and serve as a stepping stone, we make our code publicly available at https://www.github.com/joaquinsalas/COVID19-DataDriven-Classifier. Also, a live demonstration is available at http://imagenes.cicataqro.ipn.mx

## Notes

### Competing Interest Statement

The authors have declared no competing interest.

### Funding Statement

This work was partially funded by SIP-IPN 20201357 for Joaquin Salas.

### Author Declarations

Our Data Science study does not require IRB or equivalent oversight

